# Comparison of Elicit AI and Traditional Literature Searching in Systematic Reviews using Four Case Studies

**DOI:** 10.1101/2025.06.17.25329772

**Authors:** Oscar Lau, Su Golder

## Abstract

**Background:** Elicit AI aims to simplify and accelerate the systematic review process without compromising accuracy. However, research on Elicit’s performance is limited.

**Objectives:** To determine whether Elicit AI is a viable tool for systematic literature searches.

**Methods:** We compared the included studies in four systematic reviews to those identified searching with Elicit. We calculated sensitivity, precision and observed patterns in the performance of Elicit.

**Results:** Elicit had an average of 39.6% precision (26.7% - 46.2%) which was higher than the 7.55% average of the original reviews (0.65% - 14.7%). However, the sensitivity of Elicit was poor, averaging 37.9% (25.5% - 69.2%) compared to 93.5% (87.2% - 98.0%) in the original reviews. Elicit also identified some included studies not identified by the original searches.

**Discussion:** At the time of this evaluation, Elicit did not search with high enough sensitivity to replace traditional literature searching. However, the high precision of searching in Elicit could prove useful for preliminary searches, and the unique studies identified mean that Elicit can be used by researchers as a useful adjunct.

**Conclusion:** Whilst Elicit searches are currently not sensitive enough to replace traditional searching, Elicit is continually improving, and further evaluations should be undertaken as new developments take place.

**Key Messages:**

- AI tools, such as Elicit, have been developed to improve the efficiency of systematic review processes, including the identification of studies.
- Using four case study systematic reviews Elicit searches had a sensitivity between 25.5% and 69.2% (37.9% average) and precision between 26.7% and 46.2% (39.6% average).
- Elicit identified some unique studies that met the inclusion criteria for each of the case study systematic reviews.
- Elicit is constantly improving and developing its systems, thus independent researchers should continue to evaluate its performance.

## Background

Systematic reviews aim to answer a clear question by collating and synthesizing existing research on a specific topic (Cochrane, 2024). Searching on multiple databases and sources, identifying, selecting, and appraising studies are all part of the traditional methods of systematic reviews, one which relies on rigorous, time-consuming, and manual processes, often undertaken collaboratively as part of a team (Cochrane, 2024). Elicit AI aims to simplify and speed up this process by up to 80%, with a claimed time saved per review of 16 hours (Étienne Fortier-Dubois, 2025). Elicit allows the user to ask a research question and from there, is able to fully automate the systematic review by searching over 126 million papers, generating screening criteria, and providing a first draft of the review in a very short period of time. Traditionally, systematic reviews can take from 6-18 months (Wright, 2025). Policy makers, like NICE, structure guidelines based off of systematic reviews so there is a trade-off between quality and time (NICE, 2019). Waiting for a systematic review to be completed may have real world implications on patients who are being treated using potentially out-of-date guidelines. Alongside the speed and ease of use, Elicit claims to allow for more evidence to be considered and to avoid human bias without compromising on accuracy (Elicit, 2025).

AI is projected to contribute $15.7 trillion to the global economy by 2030 (National University, 2024) and in the UK alone, the number of AI companies has increased by 600% over the last 10 years (Hooson, 2025). As the use of AI in scientific research increases, the impact on research integrity becomes an ever more important factor to consider. Between 2003-2024, there has been an increase in over 15 000 papers published in Pubmed with the term ‘artificial intelligence’ (Woodhams, 2024). Of these, only 3 included both the terms ‘artificial intelligence’ and ‘research integrity’. This highlights that although research integrity is a crucial consideration when using AI tools like Elicit, there is a lack of literature surrounding this topic. Elicit may be able to significantly speed up the systematic review process, but at what cost to the rigour and reliability of traditional methods.

After compiling an answerable question, the next part of the systematic review process is to create comprehensive searches of multiple databases and other sources. This step is key to the quality of any systematic review. It is this stage of the review process that the present study evaluates by assessing the value of Elicit’s literature search facility.

Our objective is to compare the use of Elicit in identifying the included studies in systematic reviews compared to traditional search methods in systematic reviews.

## Methods

We selected four different systematic reviews in three different areas: public health (two reviews), pharmacology, and surgical procedure. This allows a comparison of Elicit across a range of subject areas to see how Elicit performs in each, thus improving the generalizability of our results. Two of the systematic reviews were conducted in 2024 by one of the authors with the searches undertaken by an experienced information specialist. The other two were also recent systematic reviews and were selected from the Cochrane Database of Systematic Reviews as Cochrane Reviews tend to be of higher quality than those published in peer review journals (Collier et al., 2006) (Windsor et al., 2012) (Fleming et al., 2012) (Moher et al., 2007) (Wen et al., 2008) (Delaney et al., 2007).

Elicit uses Semantic Scholar to find publications (Kung, 2023), searching over 126 million papers. We translated the original research question from each systematic review based on its PICO elements into an Elicit query. Next, Elicit finds the 500 most relevant studies based on the query, and the title and abstract. After this, Elicit generates automated screening criteria for these 500 studies. Some of the initial criteria did not align with those in the review, so Elicit makes it easy for us to adapt and specify the screening criteria - this does not change the 500 potentially eligible studies. Once Elicit identified the eligible studies, we exported all 500 studies into Microsoft Excel and compared the studies found in Elicit to those included in the review. Elicit makes the screening process very transparent as it provides a spreadsheet with individual columns for every inclusion criterion and if each of the studies meets it. Finally, Elicit generates a score based on the number of criteria met by the study, factoring in the importance of each one. This score is then used to determine the overall outcome of the study. Elicit stresses the fact that although it can fully automate the search, at any point in time, the user is able to ‘manually override any disagreements.’ (Byun, 2025). We noted three categories: studies that Elicit did not find, studies that Elicit found but excluded based on not meeting the screening criteria, and studies that Elicit found and included. We emailed the authors of the original review with a list of studies identified by Elicit that hadn’t been included in the original review. With the help of the authors, we were able to assess whether any of these studies met the original inclusion criteria – representing unique additional findings. We calculated the sensitivity and precision of each search using the following formulae: Sensitivity – Number of included records retrieved/Total number of included records *100, Precision – Number of included records retrieved/Total number of records retrieved *100 (Golder & Loke, 2011).

### Case Study 1: Vaping Harms

This was an umbrella review on the acute and long-term harms of vaping in young people under the age of 25 (Golder et al., in press). The authors of the review searched the KSR Evidence database on OVID, with no language restrictions on the search strategy. They also conducted a separate search for umbrella reviews on MEDLINE, Embase, and PsycINFO as these types of review are not indexed in the KSR Evidence database. The search strategy was limited to a publication date of 2015 onwards, reflecting the shift in the type of e-cigarette device being used by young people at that time. The search strategy was last conducted on the 21^st^ November 2024. In total, 381 records were screened at title and abstract stage with 56 reviews included in the final umbrella review. Thus, the precision of the searches was 56/381, 14.7%, with a Number Need to Read of 7.

For Elicit, the question asked was “What systematic reviews exist that comprehensively document the short-term and long-term health risks associated with vaping in populations aged 10-24 years?” We asked Elicit the question on the 24^th^ February 2025.

### Case Study 2: Physical Activity

This was a scoping review mapping the evidence from economic analyses on the cost-effectiveness of population-based interventions that could be funded or provided by local authorities in the UK to increase physical activity and to inform decision making at local and national level (Golder et al., 2025). The search was focused on economic literature from 2015 onwards, and contained two main segments: physical activity public health interventions in the UK and the study design being economic analyses. In total 4868 records were screened at title and abstract with 50 studies included in the final scoping review. Thus, the precision of the searches was 50/4868, 1.03%, with a Number Need to Read of 97.

For Elicit, the question asked was “What is the cost-effectiveness of population-based interventions that could be funded or provided by local authorities in the UK to increase physical activity, based on economic analyses published since 2015.” We asked Elicit the question on the 11^th^ March 2025.

### Case Study 3: Fertility Treatment

This was a systematic review published in the Cochrane Library evaluating the effectiveness and safety of vasodilators in women undergoing fertility treatment (Gutarra-Vilchez et al., 2025). The inclusion criteria consisted of: population - women of any age undergoing fertility treatment, intervention - vasodilators administered via any route with or without other agents, comparator placebo/no treatment/other active intervention, and the primary outcomes being live birth or ongoing pregnancy and the side effects of vasodilators. In total, 347 records were screened at title and abstract stage with 48 publications representing 45 studies in the final systematic review. Thus, the precision of the searches was 48/347, 13.8%, with a Number Need to Read of 7.

For Elicit, the question asked was “Evaluate the effectiveness and safety of vasodilators in women undergoing fertility treatment. Outcomes including (endometrial thickness, adverse drug reactions, live births, multiple pregnancy, ectopic pregnancy, clinical pregnancy, and miscarriage.” We asked Elicit the question on the 7^th^ March 2025.

### Case Study 4: Breast Reconstruction

This was a systematic review published in the Cochrane Library assessing ‘the effects of implants vs autologous tissue flaps for postmastectomy breast reconstruction on women’s quality of life, satisfaction, and short- and long-term surgical complications’ (Rocco et al., 2024). The inclusion criteria consisted of women undergoing primary breast reconstruction after mastectomy for breast cancer treatment or risk reduction, implant-based reconstruction compared to any autologous-tissue reconstruction. With the primary outcomes being patient reported outcomes (BREAST-Q, BRECON-31, and EORTC QLQ BRECON-23), short- and long-term complications or oncological outcomes. In total, 6308 records were screened at title and abstract stage with 41 publications representing 35 studies in the final systematic review. Thus, the precision of the searches was 41/6308, 0.65%, with a Number Need to Read of 154.

For Elicit, the question asked was “Assess the effects of implants versus autologous tissue flaps for post mastectomy breast reconstruction on women’s quality of life, satisfaction and short- and long-term surgical complications.” We asked Elicit the question on the 6^th^ March 2025.

## Results

### Case Study 1: Vaping harms

Elicit identified 38 reviews that met the screening criteria according to Elicit. Of the 38, 14 were already included in the original umbrella review. 22 studies identified by Elicit did not meet the inclusion criteria for the umbrella review. One was a scoping review (Javed et al., 2022), four were protocols of reviews (Goel et al., 2024) (MacDonald et al., 2016) (Gardner et al., 2022) (Kaltabanis et al., 2017), four were not systematic reviews (Weni Nur Aisyah et al., 2024) (Banks, Yazidjoglou & Joshy, 2023) (Tobore, 2019) (Rahim et al., 2024), six weren’t looking at the harms (Yan et al., 2023) (Mohapatra, Wisidagama, & Schifano, 2024) (Hassanein et al., 2022) (Mylocopos et al., 2023) (Gardner et al., 2024) (Lee et al., 2020), three had no analysis of young people (Arya Marganda Simanjuntak et al., 2024) (Sharma & Verma, 2020) (Glasser et al., 2017), one was a pre-print (Khouja et al., 2020a)of a study that was already included in the umbrella review (Khouja et al., 2020b) and three weren’t available in English language (Cheon et al., 2024) (Neves et al., 2024) (Araújo et al., 2024). A further two studies (Piras et al., 2020) (Becker et al., 2020a) that were identified by Elicit met the inclusion criteria of the umbrella review and would have been included if they had been identified. Both reviews were not indexed on the KSR Evidence database at the time of searching, one was a report, and the other was not indexed on MEDLINE or Embase.

Whilst Elicit included 14 of the originally included reviews, 42 of the reviews included by Golder were not included. 31 were not identified, and a further 11 were excluded based on multiple reasons such as ‘failure to meet health outcomes requirement’ (Dautzenberg et al., 2023) (Aladeokin & Haighton, 2019) and ‘uncertainty regarding specific age range.’ (Bravo-Gutiérrez et al., 2021) (Baenziger et al., 2021) (Bourke, Sharif, & Narayan, 2021) (Hua & Talbot, 2016).

The search in Elicit had a sensitivity of 27.6% (16/58) and a precision of 42.1% (16/38).

**Figure 1:**
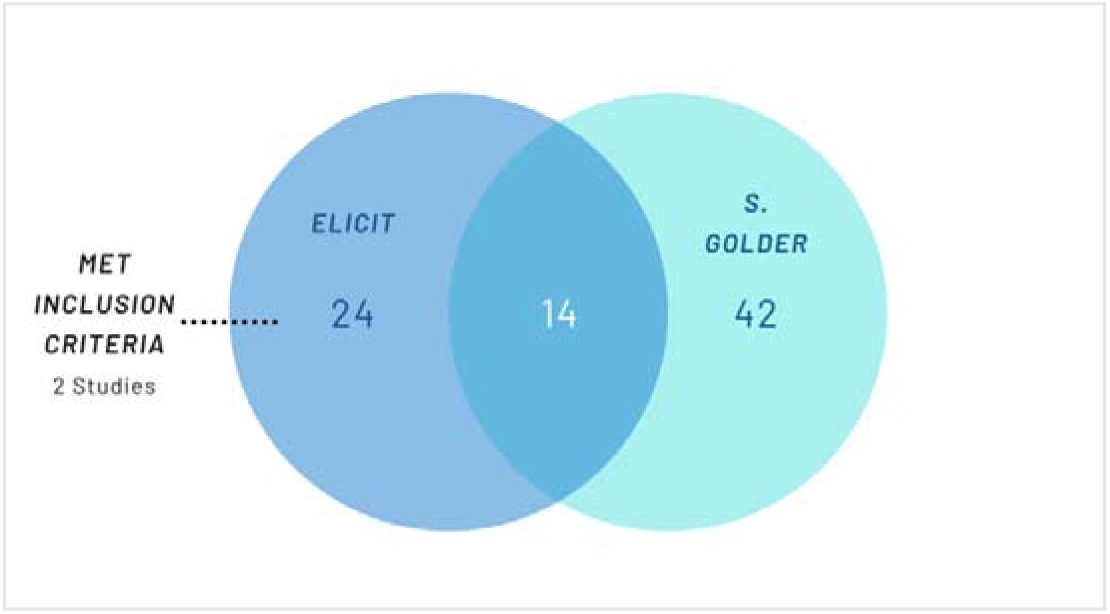
Included Studies in Case Study 1.

### Case Study 2: Physical Activity

Elicit identified 30 reviews that met the screening criteria according to Elicit. Of the 30, 12 were also included in the original scoping review (Golder et al., 2025). However, nine reviews considered eligible by Elicit were published before 2015 even though there was explicit screening criteria of a post 2015 publication date. The other eight studies identified by Elicit did not meet the inclusion criteria. Four were protocols (Mansfield et al., 2015) (Audrey et al., 2015) (Brown et al., 2017) (Howlett et al., 2017), two were not an evaluation of an intervention (Tainio et al., 2017) (Grellier et al., 2024), one did not include separate outcome data for physical activity (Gravett & Mundaca, 2021) and one had no specific intervention evaluated in terms of Return on Investment (Martin, 2015). Additionally, Elicit found one review (Aldred, Woodcock, & Goodman, 2021) that would have been included in the original review had it been identified by the traditional searches. This study was published in the Journal of Transport and Health which was not indexed in the majority of the databases searched such as MEDLINE or Embase, however, it was indexed in PsycINFO but was not identified using the economic search filter used (although it was identified by the other facets of the search).

Elicit did not include 38 of the reviews included by Golder – 27 were not identified and a further 11 were excluded based on multiple reasons such as ‘limited by the individual level approach’ (Audrey et al., 2019) (Snowsill et al.,2022) (Hunter et al., 2019), ‘narrow employee population and workplace setting limit’ (Hunter et al., 2018), ‘uncertainties around explicit UK context’ (Pretty & Barton, 2020) (Candio et al., 2021) (Jago et al., 2021) (Gc et al., 2019) and ‘significant uncertainty regarding publication year’ (Le Gouais et al., 2021) (Galbraith et al., 2021) (Kalita et al., 2022).

The search in Elicit had a sensitivity of 25.5% (13/51) and a precision of 43.3% (13/30).

**Figure 2:**
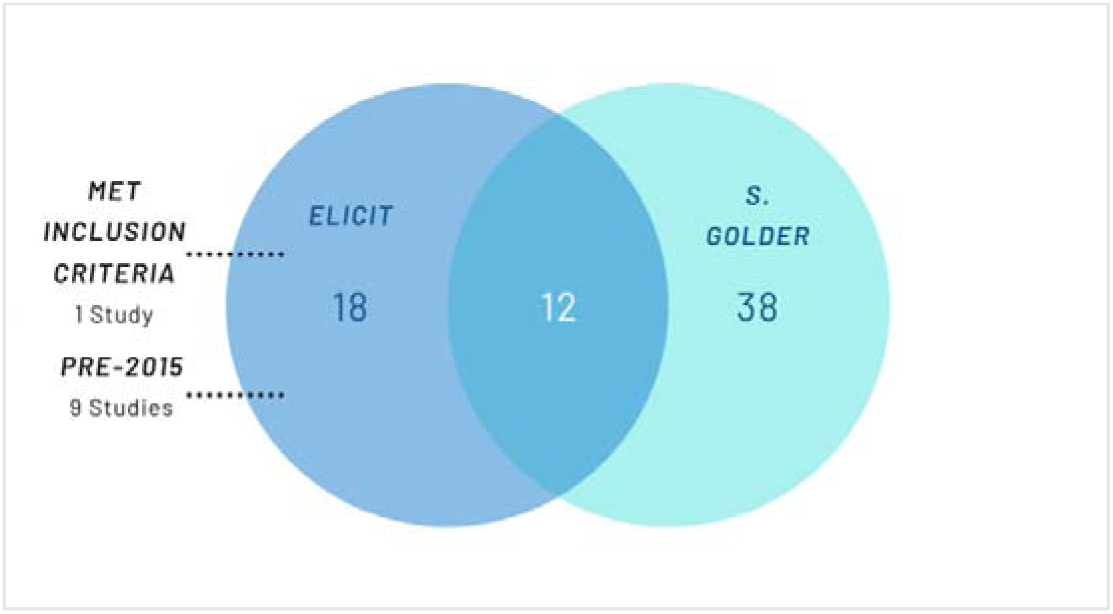
Included Studies in Case Study 2.

### Case Study 3: Fertility Treatment

Elicit identified 78 studies that met the screening criteria according to Elicit. Of the 78, 32 were included in the original Cochrane Review. Similarly, to case study 1 and 2, Elicit identified four additional studies that would have been included had they been identified at the time of searching (waleed Abd El-ghany, Ahmed, & Ayman AbdAllah Elashmawy, 2022) (Mohammed El-Khaldy et al., 2019) (Taher, Kzar Al-Essami, & Hussein Kadhim, 2023) (El-Maghrabi et al., 2020). Notably, all four RCTs found by Elicit alone were not indexed in either MEDLINE or Embase. The rest of the studies did not meet the inclusion criteria due to multiple reasons such as not being an RCT (Reddy et al., 2016) (Guo et al., 2022).

Elicit did not include 16 of the RCTs included in the original Cochrane Review– 12 were not identified and a further four were excluded for multiple reasons such as ‘significant uncertainty exists regarding specific outcomes reported and the precise study design’ (Aboelroose et al., 2020), ‘ doesn’t specifically breakdown types of pregnancy.’ (Mahran et al., 2016), ‘ambiguity about whether this fully aligns with the purposes of fertility treatment.’ (Doaa El Faham et al., 2019) and ‘doesn’t explicitly report endometrial thickness, live births, adverse reactions, ectopic pregnancy, miscarriage or multiple pregnancy.’ (Farnoush Farzi et al., 2005).

The search in Elicit had a sensitivity of 69.2% (36/52) and a precision of 46.2% (36/78).

**Figure 3:**
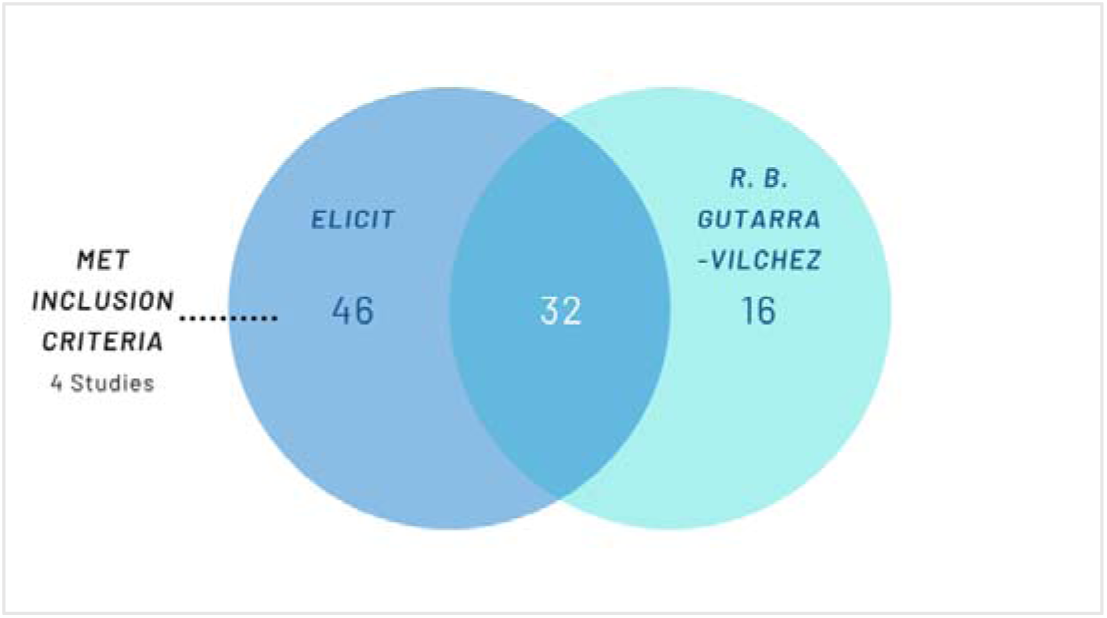
Included Studies in Case Study 3.

### Case Study 4: Breast Reconstruction

Elicit identified 45 studies that met the screening criteria according to Elicit. Of the 45, 12 were also included in the original Cochrane review. Elicit did not include 29 of the studies included in the Cochrane review – 21 were not identified and a further eight were excluded for multiple reasons such as ‘absence of patient centred outcomes’ (Ha et al., 2020), ‘ minor uncertainty regarding direct comparison between reconstruction techniques’ (Timman et al., 2017), ‘ primarily focusing on costs and technical complications’ (Aliu et al., 2017), ‘ cross sectional study’ (Kouwenberg et al., 2020), ‘quality of life measures and patient satisfaction were not measured’ (Mak & Kwong, 2020), ‘ uncertainty regarding primary vs secondary reconstruction and patient reported measures’ (Mioton et al., 2013) and ‘some population details remain implicit.’ (Xu et al., 2018) However, (Qin et al., 2018) - one of the eight excluded studies - met all the inclusion criteria but wasn’t included because the Elicit score was not high enough (it scored 4.8 with the cut-off set at greater than 4.8).

The search in Elicit had a sensitivity of 29.3% (12/41) and a precision of 26.7% (12/45).

**Figure 4:**
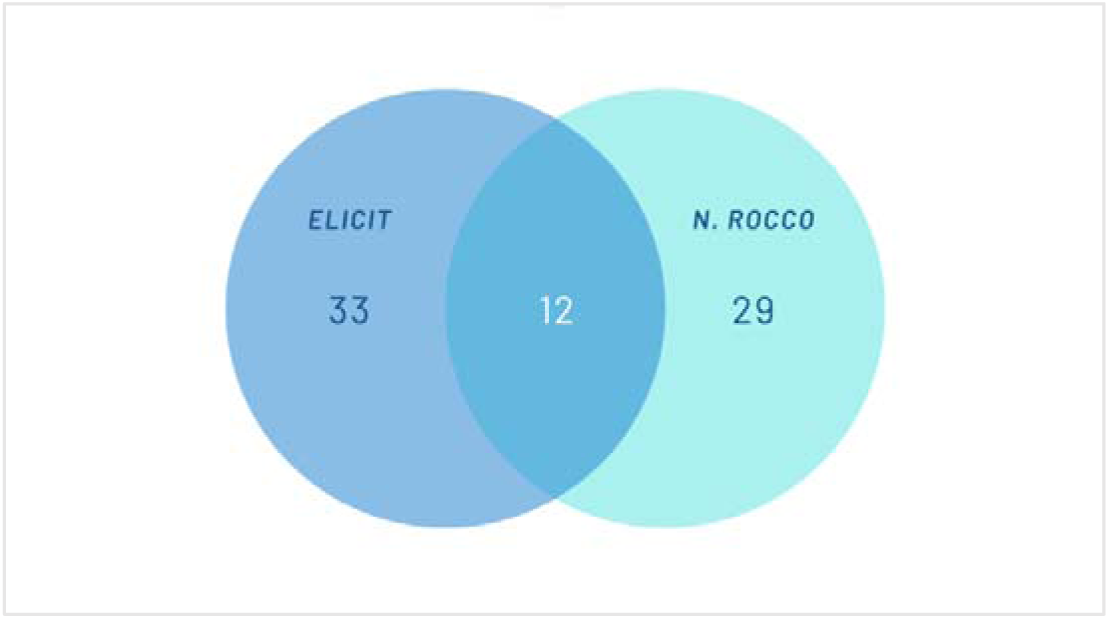
Included Studies in Case Study 4.

**Table 1:** Studies Identified by Elicit and Review with Traditional Searching Methods for Each Case Study.

|  | Elicit Inclusion | Included in Traditional Review | Not Included in Traditional Review and Did Not Meet Inclusion Criteria | Not Included in Traditional Review but Did Meet Inclusion Criteria | Elicit Precision (%) | Elicit Sensitivity (%) | Traditional Searching Precision (%) | Traditional Searching Sensitivity (%) |
| --- | --- | --- | --- | --- | --- | --- | --- | --- |
| Case Study 1: Vaping harms | 38 | 14 | 22 | 2 | 16/38 (42.1%) | 16/58 (27.6%) | 56/381 (14.7%) | 56/58 (96.6%) |
| Case Study 2: Physical Activity | 30 | 12 | 17 (9 pre-2015) | 1 | 13/30 (43.3%) | 13/51 (25.5%) | 50/4868 (1.03%) | 50/51 (98.0%) |
| Case Study 3: Fertility Treatment | 78 | 32 | 42 | 4 | 36/78 (46.2%) | 36/52 (69.2%) | 48/347 (13.8%) | 48/52 (92.3%) |
| Case Study 4: Breast Reconstruction | 45 | 12 | 33 | ? (waiting for response) | 12/45 (26.7%) | 12/41 (29.3%) | 41/6308 (0.65%) | ? Waiting |

**Figure 5:**
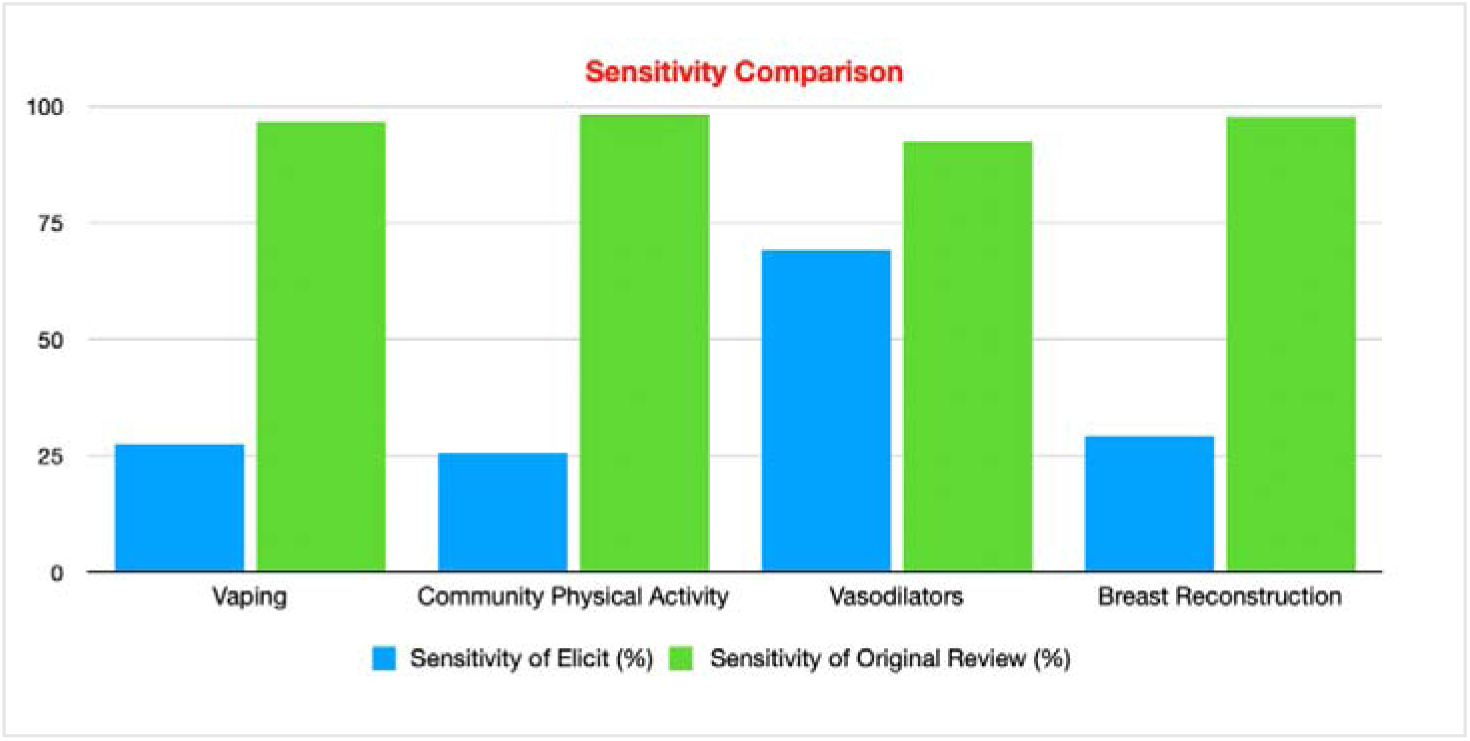
Sensitivity Comparison of Elicit and Traditional Review for Each Case Study.

**Figure 6:**
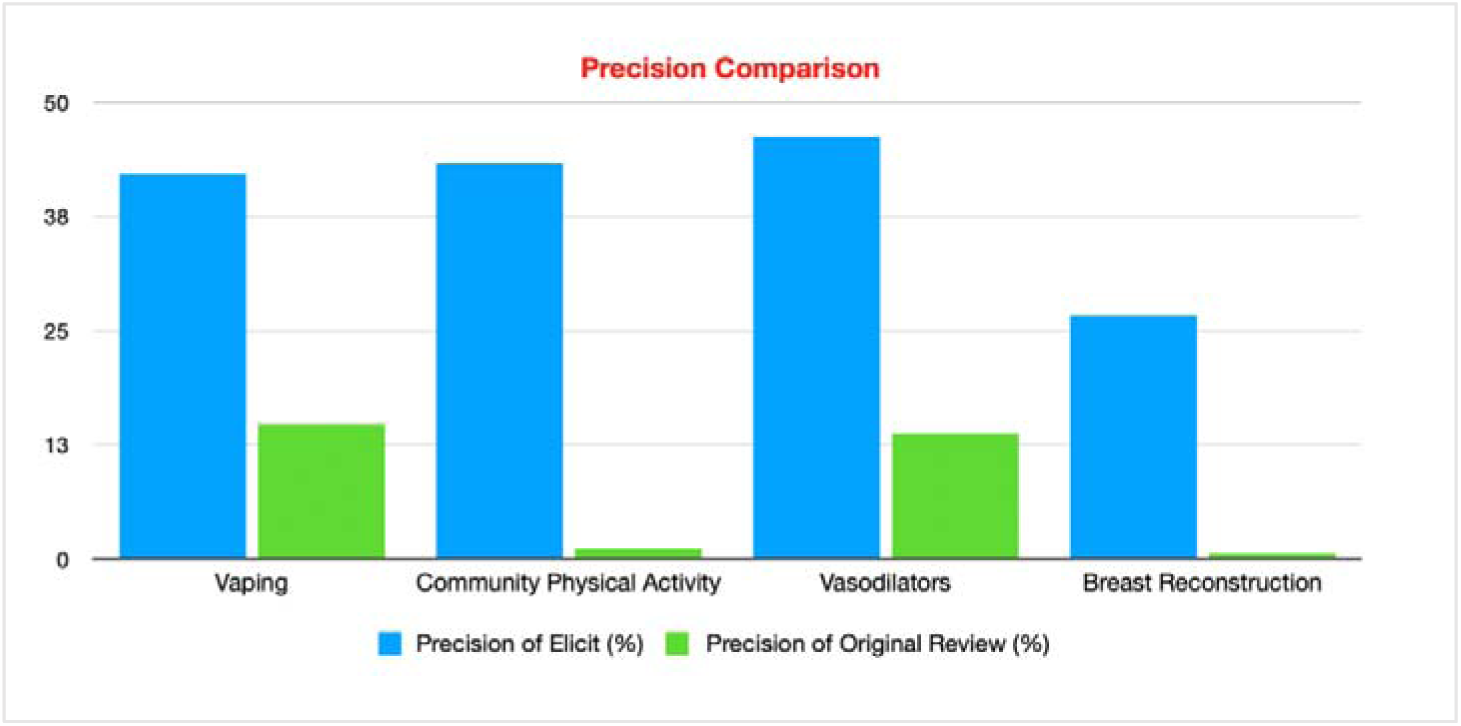
Precision Comparison of Elicit and Traditional Review for Each Case Study.

## Discussion

The primary aim of this study was to assess Elicit’s ability to identify studies that meet the inclusion criteria for a systematic review. We hypothesized that Elicit may be able to assist in the literature search and screening of studies as part of the systematic review process, and with improvements even replace traditional searching methods. We also sought to determine whether Elicit is comparable to traditional searching in terms of recall and precision.

Across case studies 1,2 and 3, Elicit was able to find and include additional studies that the original authors didn’t. This may be due to Elicit’s ability to search on a wider set of records (Elicit, 2025), as it uses Semantic Scholar which indexes over 126 million articles. This, coupled with Elicit’s speed and high precision makes it useful for conducting preliminary searches for example, costing a grant proposal, providing seed papers for improving traditional searches or testing search strategies, or determining whether there is a risk of an empty review. The fact that Elicit provides detail for each of the studies and reasoning for inclusion/exclusion in the screening process is an advantage over many other AI tools and it also allows the user to understand to some extent what Elicit is doing in real time and make adjustments - strengthening the research integrity (Woodhams, 2024). Whilst Elicit may not be at the stage of development to replace traditional searching methods, Elicit’s ability to assist in the search process is very apparent through its speed and precision. Precision using Elicit across the four systematic reviews had an average of 39.6% which is impressively high, compared to the average of the original reviews which was 7.55%. With such high precision, Elicit would be invaluable in the costing and proposal stage of a review.

Little independent research on the performance of Elicit has been conducted to date. Fenske and Otts (2024) conducted a descriptive study comparing Elicit to Pubmed and CINAHL. They surveyed 323 graduate nursing students, with the primary outcome being which resource they preferred and their opinions on Elicit. As the study was conducted in the fall of 2023, they used a beta version of Elicit. Of the 26% of students who preferred Elicit, 38.8% listed the user-friendliness as a top strength. This aligns with our study as Elicit has a very shallow learning curve which allows for it to be easily implemented as an auxiliary tool alongside a traditional literature search. By using Elicit as an adjunct, as shown in our case studies, it may allow the researcher to find additional studies that otherwise wouldn’t have been identified.

However, in our study we noticed some key flaws in Elicit. For case study 2, Elicit deemed nine studies-which were published before 2015-eligible even though we explicitly added to the screening criteria to only include publications from 2015. This shows that even through manually adjusting screening criteria, Elicit is not perfect and studies can slip through the cracks in the screening stage. This potentially contradicts the benefit of being able to change the criteria by hand. Raising the question of the value of being able to make adjustments if Elicit doesn’t always apply the criteria. The sensitivity for systematic review searches should reach at least 90% (Beynon et al., 2013). However, due to Elicit’s limited sensitivity - average of 37.9% - this means that it currently cannot be used as a one stop shop in systematic review searches. Out of the four case studies, case study 3 (Gutarra-Vilchez et al., 2025) had by far the highest sensitivity of 69.2%. A potential reason for this may be because the PICO structure and inclusion criteria for pharmacology reviews are more straight forward, well-defined and binary. Alternatively, the two public health reviews had the lowest sensitivity, highlighting potential limitations with Elicit when the PICO is more ambiguous and complex.

Furthermore, in case study 2, Elicit identified a pair of duplicate studies (Charles et al., 2019). However, one of the duplicates was included and the other one was excluded. This questions whether Elicit is effective in de-duplication (Darowski, 2023) when the study appears in different sources. Furthermore, if one copy was included and the other excluded, this appears to show possible inconsistencies in the screening process of Elicit which may lead to an overestimation in the number of included studies.

Interestingly, in case study 4, the Elicit score threshold (as mentioned previously) was set at greater than 4.8, with (Qin et al., 2018) scoring 4.8, thus being excluded. However, several other studies - for example (Hu et al., 2009) (Yueh et al., 2010) (Kamel et al., 2019) - also had a score of 4.8 but were included by Elicit. This highlights potential inconsistencies in Elicit’s screening, proving that human oversight remains an integral part when interpreting borderline cases. Further investigation into the scoring process may explain why studies with identical scores receive different outcomes and whether supplementary criteria are affecting inclusion.

A recently published study by Bernard et al. (2025) evaluated the repeatability, reliability, and accuracy of Elicit in one case study. The authors searched Elicit on the 18^th of^ April 2023 and had similar findings to our study. Elicit identified three unique studies that weren’t included in the umbrella review but did meet the inclusion criteria. The ability of Elicit to find studies that otherwise wouldn’t have been included is a strong argument for Elicit as an adjunct. However, just like our study, Bernard et al found that the sensitivity of Elicit is poor. As it was only able to find 3 out of 17 studies (17.6%) included in the umbrella review. In our case studies using the same methods of calculation as Bernard et al, Elicit found 25.0%, 24.0%, 66.7% & 29.3% respectively. Bernard et al. conducted their evaluation two years ago (April 2023), demonstrating that Elicit has made progress, but is still unable to replace a traditional literature search.

Building on from both our study and Bernard et al., Tomczyk et al. (2024) evaluated the search ability of Elicit against those of experienced researchers using Scopus and Web of Science, focusing on nine marketing and e-commerce themed research questions. Their findings were consistent with ours, as they concluded that Elicit demonstrated “great potential in surfacing rare literature that other means miss.” Suggesting that the key question is not whether Elicit can replace traditional search methods, but how it can enhance systematic review processes by complementing conventional approaches.

There are a number of limitations of our study. The nature of our study was retrospective as we were analysing Elicit after the systematic reviews had been carried out. Although we did aim to take into account the difference in timing of the searches using Elicit and the traditional searches, this was not always possible. Additionally, Elicit is being constantly updated as “if you asked once before the update and once after, you might get two different answers from two different version of Elicit.” (Elicit, 2024) This will be a limitation of any evaluation of the performance of Elicit, as the results and conclusions may be different as new versions of Elicit are released.

Strengths of our study include the use of four reviews in different areas of medicine which enhanced the generalizability and robustness of our findings. We were also able to identify Elicit’s strengths and limitations, and spot patterns with how Elicit works for different topics. Furthermore, there is a lack of prior research on Elicit for systematic searches. So, this study is important in quantifying the sensitivity and precision of Elicit which can help in deciding if Elicit currently is a viable research tool.

## Conclusions

This study highlights the high precision of the current version of Elicit AI, making it useful for writing grant proposals, and conducting scoping and preliminary searches. The unique studies identified in Elicit demonstrate that it can be a useful adjunct to traditional searching, but it is not equipped to replace standard systematic searching due to its low sensitivity across the four reviews. Further independent evaluations of Elicit are needed as Elicit is updated and newer versions are released.

## Data Availability

All data produced in the present study are available upon reasonable request to the authors

## Acknowledgements

We would like to thank the authors of case study 3 and 4 for their invaluable contributions to screening the new records identified by Elicit.

## Ethical Approval

Ethical approval was not required for this project, as only public available secondary data was used.

